# Coupled stack-up volume RF coils for low-field open MR imaging

**DOI:** 10.1101/2024.08.30.24312851

**Authors:** Yunkun Zhao, Aditya A Bhosale, Xiaoliang Zhang

## Abstract

**Background:** Low-field open magnetic resonance imaging (MRI) systems, typically operating at magnetic field strengths below 1 Tesla, has greatly expanded the accessibility of MRI technology to meet a wide range of patient needs. However, the inherent challenges of low-field MRI, such as limited signal-to-noise ratios and limited availability of dedicated radiofrequency (RF) coils, have prompted the need for innovative coil designs that can improve imaging quality and diagnostic capabilities.

**Purpose:** In response to these challenges, we introduce the coupled stack-up volume coil, a novel RF coil design that addresses the shortcomings of conventional birdcage in the context of low-field open MRI.

**Methods:** The proposed coupled stack-up volume coil design utilizes a unique architecture that optimizes both transmit/receive efficiency and RF field homogeneity and offers the advantage of a simple design and construction, making it a practical and feasible solution for low-field MRI applications. This paper presents a comprehensive exploration of the theoretical framework, design considerations, and experimental validation of this innovative coil design.

**Results:** We demonstrate the superior performance of the coupled stack-up volume coil in achieving 47.7% higher transmit/receive efficiency and 68% more uniform magnetic field distribution compared to traditional birdcage coils in electromagnetic simulations. Bench tests results show that the B1 field efficiency of coupled stack-up volume coil is 57.3% higher compared with that of conventional birdcage coil.

**Conclusions:** The proposed coupled stack-up volume coil outperforms the conventional birdcage coil in terms of B1 efficiency, imaging coverage, and low-frequency operation capability. This design provides a robust and simple solution to low-field MR RF coil design.

## 1. Introduction

Magnetic resonance imaging (MRI) (1,2) has evolved into an indispensable tool for clinical diagnosis and basic biomedical research (3–5), offering non-invasive and high-resolution visualization of anatomical structures (6–9), physiological processes (10–12), and functional (13–15) and metabolic (16–19) information within the human body. While high-field MRI (20–22) has demonstrated a significant SNR gain (23–25) and dominated the field (26–28), low-field MRI (below 1 Tesla) (29–32) has garnered significant attention in recent years due to its unique advantages and clinical utility (33,34), as well as recent advances in artificial intelligence (35–38). The appeal of low-field open MRI lies in its capacity to cater to a diverse patient population, including those with claustrophobia, obesity, and pediatric patients, who may find conventional closed-bore MRI systems challenging or uncomfortable (39).

Low-field MRI systems, characterized by magnetic field strengths below 1 Tesla, have gained significant attention due to their affordability, improved safety profile, and increased accessibility (40). However, the shift to lower magnetic field strengths introduces challenges, particularly the inherently lower signal-to-noise ratio (SNR) (41,42). This reduction in SNR can compromise image resolution and hinder the detection of subtle anatomical or pathological details, underscoring the need for innovative solutions to maintain high imaging quality in low-field MRI.

Open MRI systems are generally low-field systems, offering a spacious, open architecture that enhances patient comfort and accessibility. These systems are especially beneficial for certain patient populations, including those with claustrophobia, obesity, or pediatric patients, who may find conventional closed-bore MRI systems challenging. Additionally, the open design facilitates a broader range of imaging scenarios, such as interventional procedures and imaging of larger anatomical regions, making them a versatile tool in clinical practice. However, while these benefits are significant, they also introduce specific challenges that must be addressed, particularly in the context of maintaining image quality at lower field strengths.

Central to the success of any MRI system is the radiofrequency (RF) coil (43–45), a crucial component responsible for transmitting and receiving MR signals during the imaging process. The design and performance of the RF coil play a pivotal role in image quality, signal strength, noise level, and overall diagnostic accuracy (46–49). In the context of low-field open MRI, the current RF coil configurations face the challenge of limited RF field (B1 field) transmit/receive efficiency and field homogeneity, particularly along coil axis. To bridge this gap and harness the full potential of low-field open MRI systems, we introduce a coupled stack-up volume coil, a novel RF coil design specifically tailored for head imaging at a Larmor frequency of 21.3 MHz to address these challenges. Previous designs of stack-up coils have primarily focused on achieving decoupling between the individual coils to minimize interference and optimize performance. In contrast, our design intentionally allows the coils to be strongly coupled with each other, which enhances the overall B1 field efficiency and homogeneity within the imaging volume. To investigate and demonstrate the proposed design, we have taken the 0.5T as an example field strength and designed and constructed a prototype coupled stack-up volume coil operating in the 20 MHz range. This RF coil design can significantly improve RF field efficiency and also enhance the field homogeneity along the coil axis (i.e. imaging coverage), ultimately elevating the performance of low-field open MRI systems. The proposed design of the coupled stack-up coil was analyzed using full-wave electromagnetic (EM) simulation and tested on the workbench with standard RF measurement procedures. The performance is further validated through a comparison study with a standard birdcage coil (46,50).

## 2. Methods

### 2.1 EM Simulation

Finite difference time-domain simulation software CST Studio Suite (Dassault Systèmes, Paris, France) was used to obtain numerical results of the proposed designs. Figure 1A shows the layout of the coupled stack-up volume coil. The coupled stack-up volume coil design consists of a stack of seven identical and individual coils, meticulously arranged to create a cylindrical imaging area with dimensions of 300 mm in diameter and 300 mm in length. Each coil unit is equipped with a 60 pF capacitance capacitor, carefully selected to optimize its resonance characteristics at the desired Larmor frequency of 21 MHz. The coil is driven via the central coil element in this stack configuration, which provides efficient RF signal transmission and reception throughout the imaging volume. The spacing between these individual coils was carefully arranged based on the observation that in uniformly spaced coil arrays, the magnetic field strength tends to be weaker at the sides of the coil compared to the center. To address this, we adjusted the gaps so that they are smaller at the sides and larger at the center, thereby enhancing the field strength at the coil’s edges. The precise gap distances were determined through a process of trial and error, involving multiple simulations and iterative adjustments, until the optimal configuration for maximum field efficiency and homogeneity was achieved. The circuit diagram and coil spacing are shown in Figure 1B. Based on the number of coils, there are four resonant modes for the coupled stack-up coil, and the lowest resonant mode is used for imaging because it has the highest field strength efficiency. A traditional 8-leg low-pass birdcage coil and a 7-turn solenoid coil, both with the same size as the coupled stack-up coil, have also been built for comparison. In a comparison study, a cylindrical oil phantom (σ(conductivity) = 0 S/m εr(relative permittivity) = 2.33, diameter = 20 cm, length = 30 cm) was placed centrally along the axis of the coils, with the entire volume of the phantom considered as the region of interest (ROI) for field strength and distribution evaluation. Scattering parameters and B1 field efficiency map were used to evaluate the performance of the stacked coils in coupling and imaging. To determine the performance of the proposed design under more realistic conditions, the coupled stack-up volume coil and birdcage coil were simulated on the human model Gustav for head imaging. Material properties of the human model at 21 MHz were taken from CST Studio Suite. All magnetic and electric field plots were normalized to 1 W accepted power, meaning that the field strength values were divided by the square root of the accepted power to ensure consistent comparison across different scenarios.

**Figure 1.**
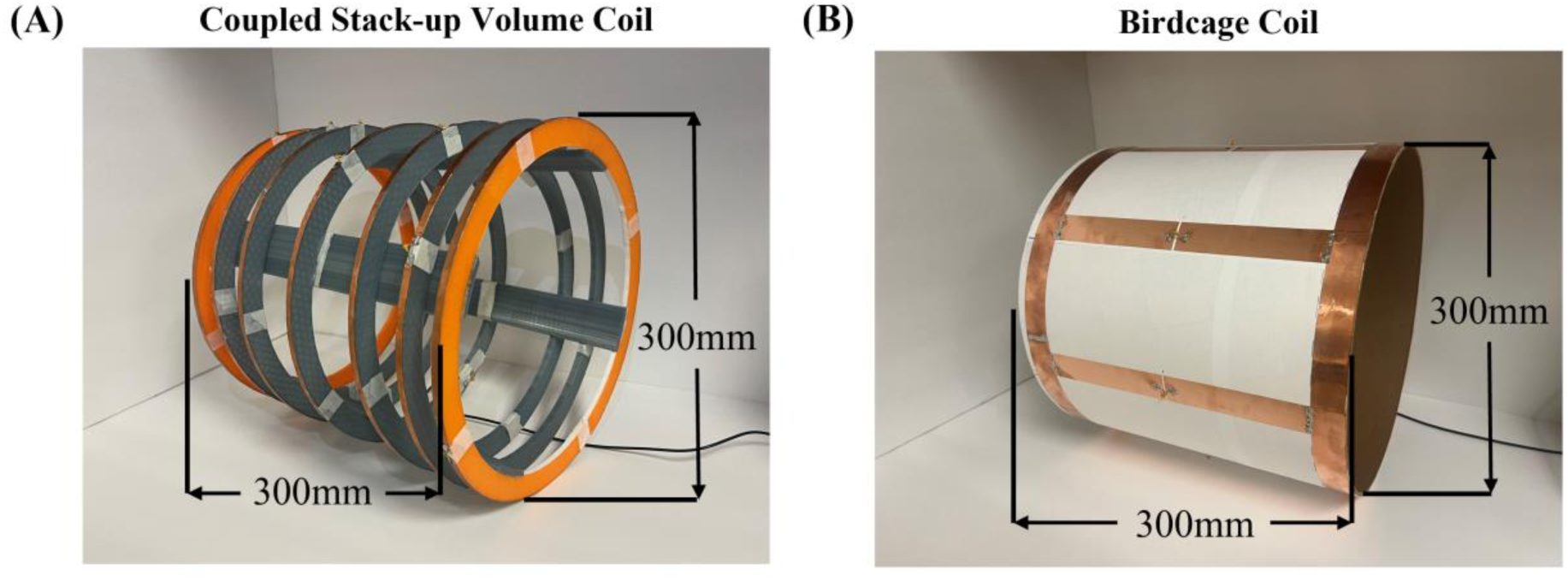
(A) Simulation model of coupled stack-up volume coil. (B) Circuit diagram of coupled stack-up volume coil. The distance between individual coils has been labeled in the figure.

### 2.2 Bench Test Model Assembly

Figure 2A shows photographs and dimensions of bench test models of the coupled stack-up volume coil and birdcage coil. The bench test models have the same dimensions as the simulation model. The electrical track of the coupled stack-up volume coil was built using 6.35 mm-wide copper tape and mounted on a 3D-printed polylactide acid frame. Due to the width of the 3D-printed frame, the inner diameter is 260 mm and 40 mm shorter than the simulation mode. The imaging resonant frequency was tuned to 21 MHz and matched to 50 ohms by careful selection of the capacitance value on each individual coil. We used 7 identical fixed tuning capacitors with 39 pF capacitance. One capacitor with 330 pF connected in parallel to the feeding line was employed for impedance matching. Except for three coils located at the center and two sides, the remaining four coils are movable, and their position may be adjusted to achieve a homogenous B-field under different imaging objects. Most areas of the coil are hollow and can also be used to alleviate claustrophobia in patients.

**Figure 2.**
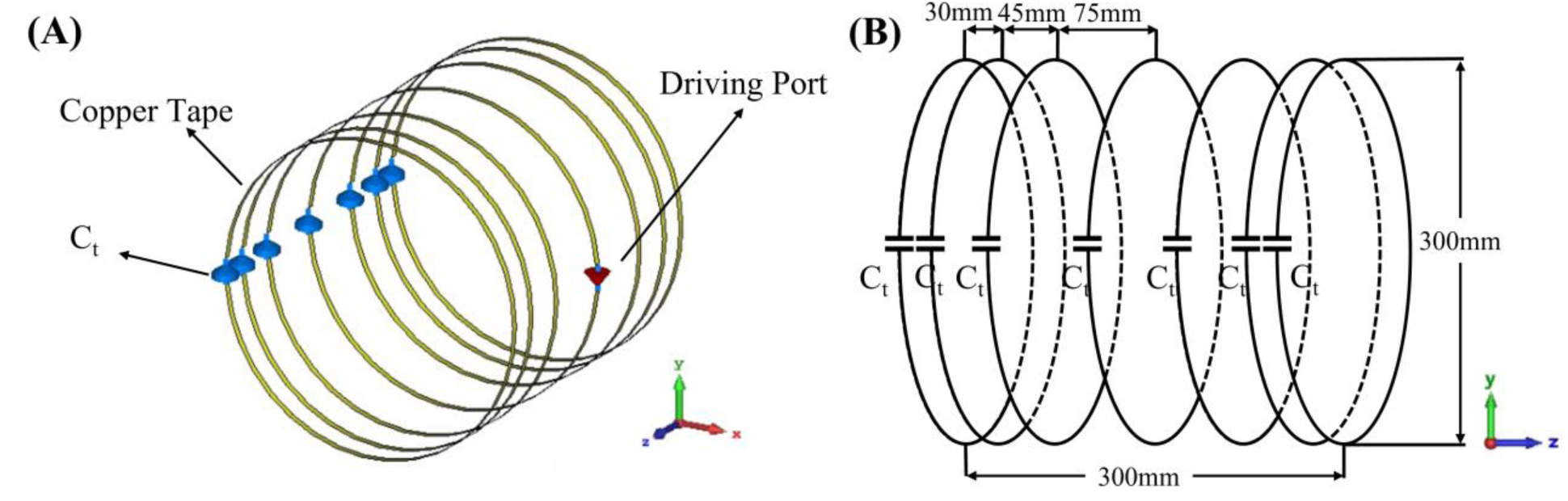
A photograph (A) of the bench test coupled stack-up volume coil model for imaging at 0.5T, corresponding resonant frequency of 21 MHz. For comparison, a custom-built 21 MHz low-pass birdcage coil (B) was used in this paper.

For comparison, a low-pass birdcage coil has also been made. The birdcage coil model has the same dimensions as its simulation model and the coupled stack-up volume coil. It was built using 6.35 mm-wide copper tape on a cardboard structure. The birdcage coil has 8 legs with 8 tuning capacitors and was tuned to 21 MHz and matched to 50 ohms by tuning capacitors and a matching circuit.

### 2.3 3-D Magnetic and Electric Field Mapping

A sniffer positioning system combined with a magnetic and electric field measurement setup, shown in Figure 3, was used to visualize the B and E field distribution in the bench test. The system consists of a Genmitsu PROVerXL 4030 router (SainSmart, Lenexa, United States) as a positioning system, a Keysight E5061 Vector Network Analyzer (Keysight, Santa Rosa, United States) for data reception and analysis, and a B/E field sniffer to receive field strength data. The positioning system was programmed to measure the B or E field strength at a level above the coils with a data step of 0.5 mm. The design of the B and E field sniffers is also shown in Figure 3. The B field sniffer is a coaxial cable loop that can measure the magnetic flux flow through the center of the loop, and the E field sniffer is a coaxial cable with the outer conductor and medium removed at the tip. During the measurement, the coil assembly is connected to port 1 of the VNA, and the sniffer is connected to port 2. The S21 value is recorded by the VNA, and the final field strength is calculated using the following equation:

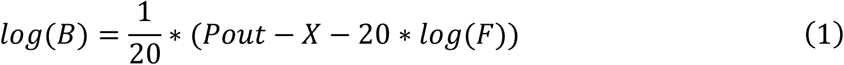

**Figure 3.**
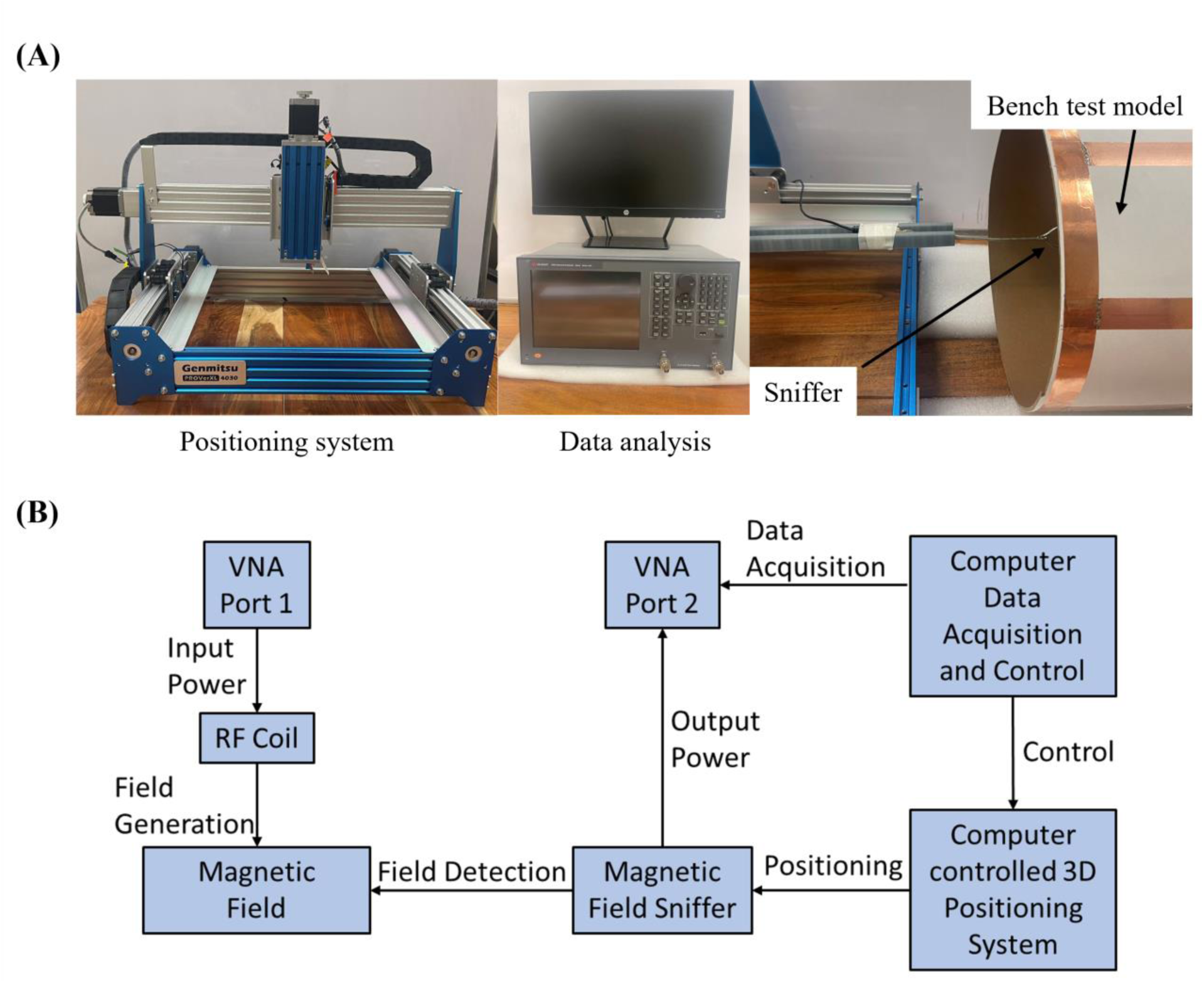
(A) Experimental setup of the sniffer-positioning system combined magnetic field measurement for the coupled stack-up volume coil. The FOV of the measuring system is 200 mm * 150 mm * 80 mm, and the resolution is 0.5 mm * 0.5 mm. (B) Data processing flow for the 3-D magnetic field mapping system.

Where *B* is the magnetic flux density in Tesla, *F* is the frequency of the received signal in megahertz, *Pout* is the probe output power into 50 ohms in dBm, and *X* is a scale factor from calibration. The calibration was taken place using the result from the magnetostatics method and finite-difference time-domain (FDTD) method on a 5cm diameter circular RF coil with one tuning and one matching capacitor and built with 16 AWG copper wire. Three calculation results were used, including the numerical solution and analytical solution of the Biot-Savart law:

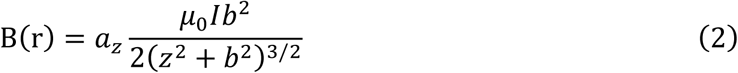

The Biot-Savart law is used to find the magnetic flux density at a point on the axis of a circular loop of radius *b* that carries a direct current *I* to verify the magnetic field. The result from FDTD methods generated by the electromagnetic simulation model from simulation software CST Studio Suite has been used to verify electric measurement results. All three calculated and simulated results verified our measurement system is correct and accurate.

## 3. Results

### 3.1 Simulated Resonant Frequency and Field Distribution

Simulated scattering parameters versus frequency of the stacked coils are shown in Figure 4A. As shown in the figure, strong coupling is created between the coils, resulting in split resonant peaks. Four resonant frequencies were generated, with the lowest frequency at 21 MHz and the highest at 37.6 MHz. Figure 4B presents the normalized field distribution for four different resonant modes of the coupled stack-up volume coil. Among these modes, only the lowest mode exhibits the strongest B1 field efficiency and a uniform field direction, making it the most suitable for MR imaging applications. The other modes show less efficient and less uniform field distributions, which are not ideal for imaging purposes. For mode 1, the unloaded Q factor is 381.41 and loaded Q is 51.06. Figure 5 shows simulated Y-Z, X-Z, and X-Y plane B field efficiency maps inside phantom generated by coupled stack-up volume coils, in which both planes are at the center of the axis. A set of the multiple X-Y plane slices with different distances from the phantom center B field efficiency maps inside the phantom generated by coupled stack-up volume coil has also been shown. The simulation result shows the coupled stack-up volume coil has great field homogeneity, which can be used for MR imaging.

**Figure 4.**
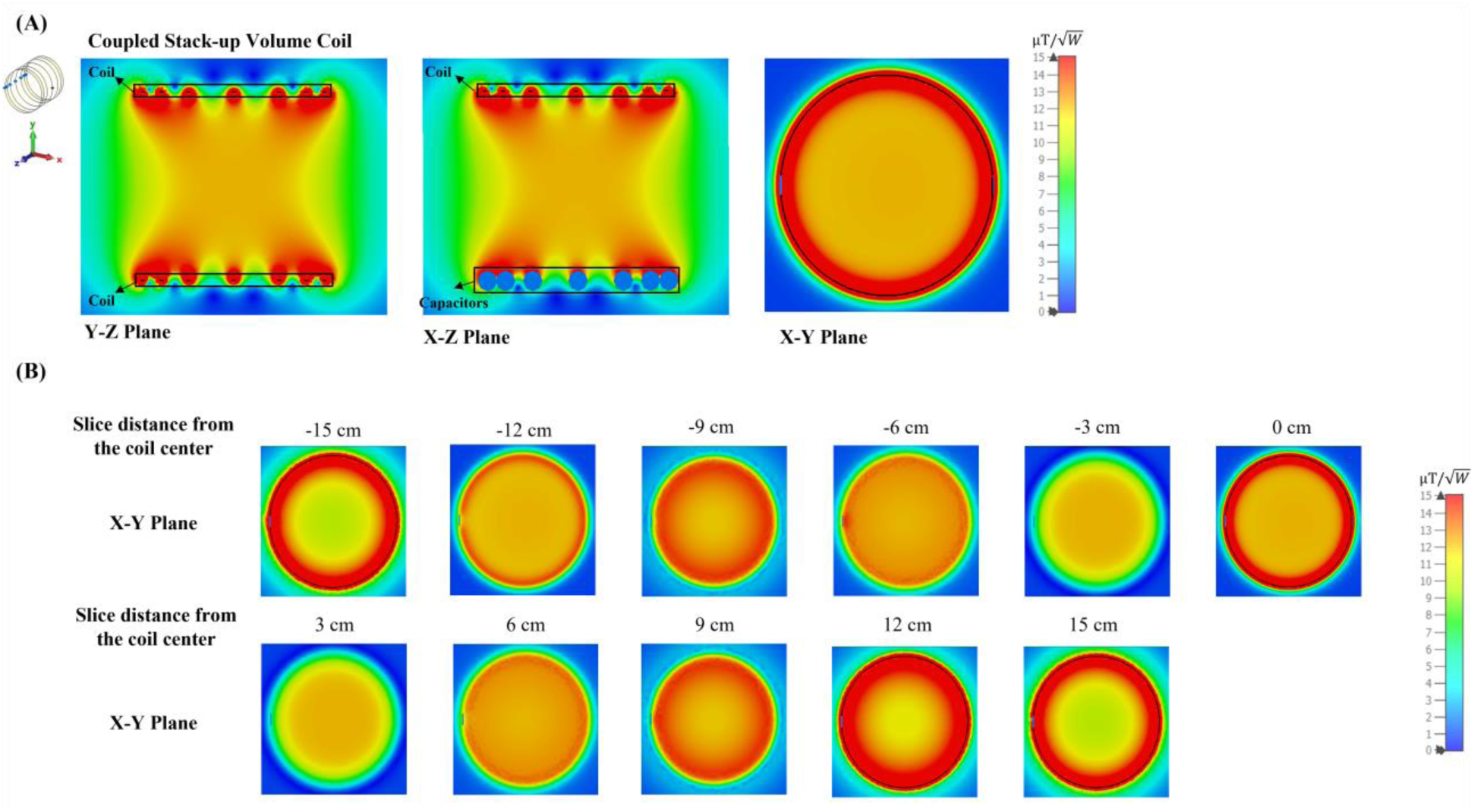
(A) Simulated S11 vs. frequency of the coupled stack-up volume coils. (B) Normalized B1 field distribution for each mode.

**Figure 5.**
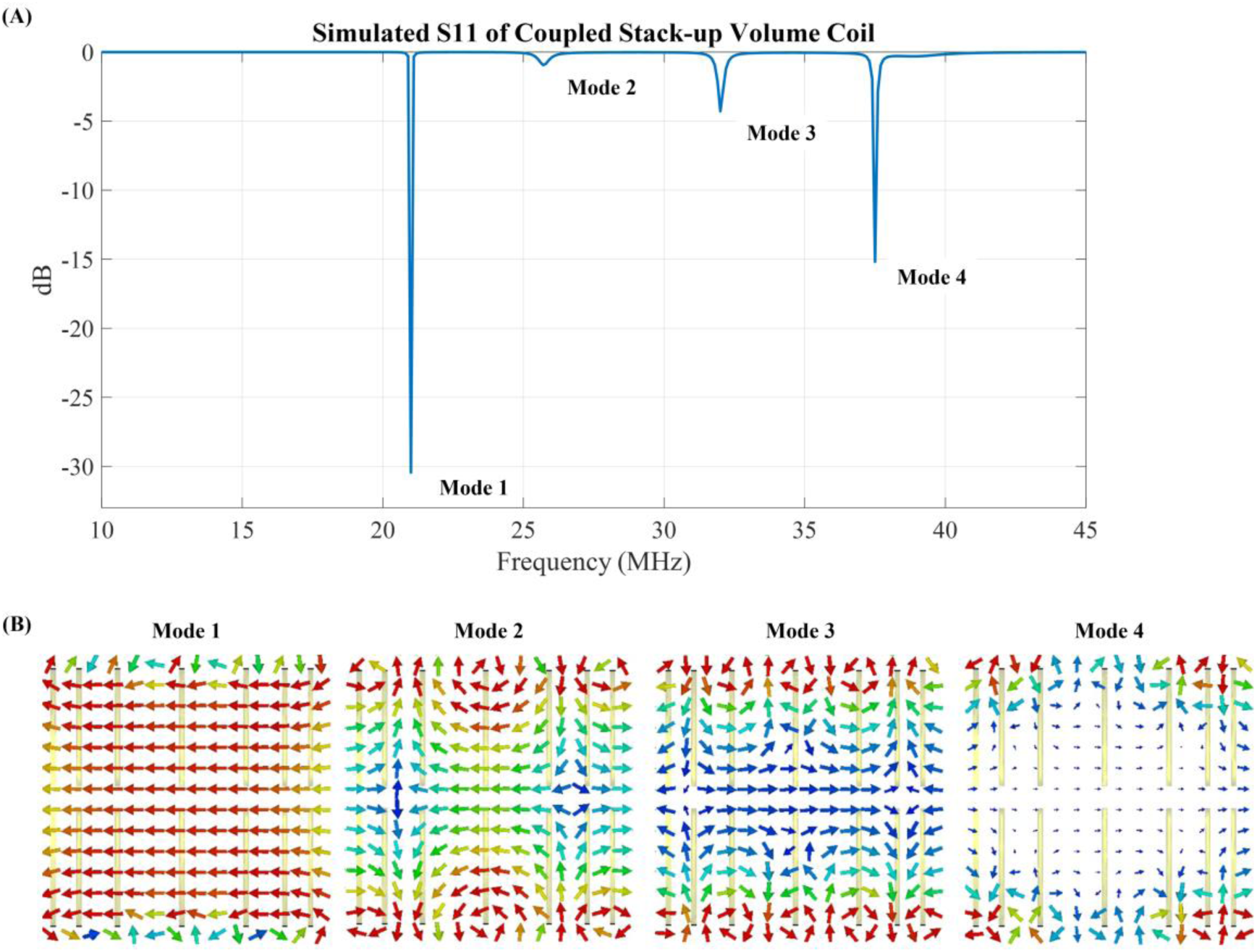
(A) Simulated unloaded Y-Z, X-Z, and X-Y plane B field efficiency maps inside oil phantom generated by coupled stack-up volume coils. Both planes are at the center of the axis. (B) A set of the multiple X-Y plane slices with different distances from the phantom center B field efficiency maps inside the phantom generated by coupled stack-up volume coil.

### 3.2 Measured Scattering Parameters and Field Distribution

Figure 6A shows that the S-parameter vs. frequency plots of the coupled stack-up coil are in good agreement with the simulation results. Four resonant modes with 20.1 MHz, 28.2 MHz, 31.8 MHz, and 34.4 MHz were formed. Figure 6B shows the B field efficiency distribution map on the Y-Z plane measured with a 3-D magnetic field mapping system. Coupled stack-up volume coil shows significant homogeneity and strong B field efficiency on the Y-Z plane and is in accordance with the simulation result, which also indicates that the simulation results are accurate and reliable.

**Figure 6.**
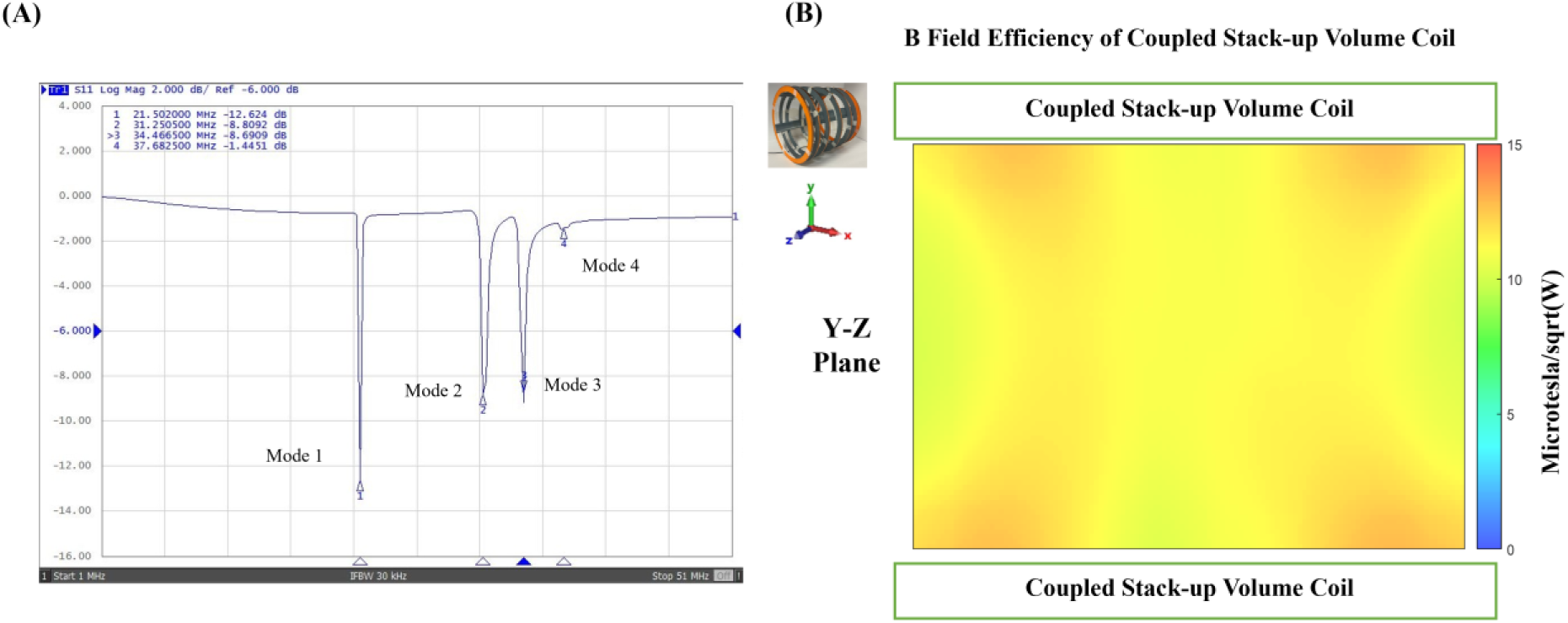
(A) Scattering parameters vs. frequency of the bench test model of coupled stack-up volume coils. (B) Measured unloaded B field efficiency maps on the Y-Z plane of coupled stack-up volume coil.

### 3.3 Field Distribution and Efficiency Evaluation

To comprehensively evaluate the performance of our proposed design, we compared three different coil setups: the coupled stack-up volume coil, the equal gap coupled coil, and a 7-turn solenoid coil. Each configuration was designed with the same dimensions to ensure a fair comparison. The coupled stack-up volume coil was designed with variable gaps between the individual coils to enhance field homogeneity, while the equal gap coupled coil features evenly spaced coils along its length. The solenoid coil, commonly used in low-field MRI due to its ability to generate a uniform magnetic field along the B1 direction, was included as a benchmark.

Figure 7A shows the simulation models of the equal gap coupled coil and the solenoid coil. In Figure 7B, the simulated B1 field efficiency comparison is presented, while Figure 7C shows the simulated E field efficiency comparison. Figure 7D provides a 1-D profile of the field efficiency along the dashed line in Figure 7B. The results indicate that the equal-gap coupled coil and the solenoid coil exhibit relatively similar B1 field efficiencies, with both showing strong efficiency at the center of the coil. However, the coupled stack-up volume coil, while slightly lower in field efficiency at the center, demonstrates superior field homogeneity across the imaging area. This advantage in homogeneity makes the coupled stack-up volume coil more adaptable for real-world applications, where varying the layout of the coils can further optimize performance.

**Figure 7.**
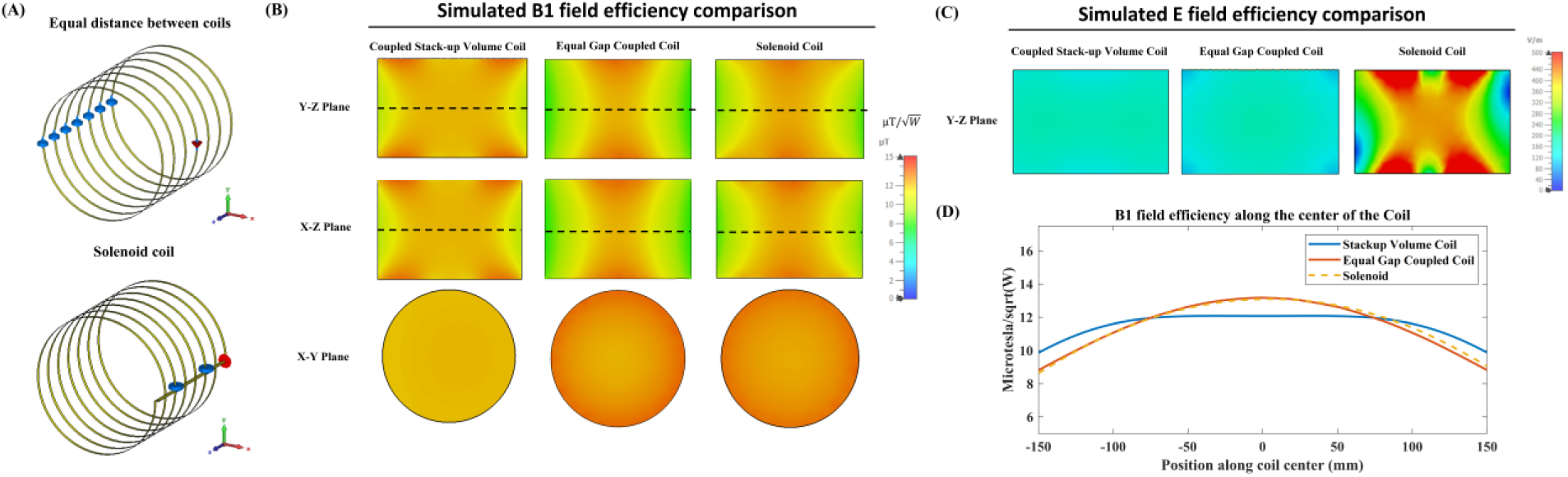
(A) Simulation models of the equal gap coupled coil and the 7-turn solenoid coil. (B) Simulated unloaded B1 field efficiency comparison among the coupled stack-up volume coil, the equal gap coupled coil, and the solenoid coil. (C) Simulated unloaded E field efficiency comparison between the three coils. (D) 1-D profile of the field efficiency along the center line of the coils (dashed line in Figure 7B), showing the field distribution along the coil axis.

In terms of E field efficiency, which generally correlates with noise in SNR calculations, the coupled stack-up volume coil significantly outperforms the solenoid coil, exhibiting much lower E field values. This suggests that the coupled coil design could potentially generate less noise in actual imaging, leading to improved image quality and overall performance in low-field MRI systems.

To further validate our findings, we conducted additional simulations with a CST Studio bio-model loaded into the coils. Figure 8A presents the simulated B1 field efficiency comparison, while Figure 8B shows the simulated E field efficiency comparison. Finally, Figure 8C illustrates the simulated specific absorption rate (SAR) comparison between the different coil setups. The results from the bio-model simulations are consistent with those obtained using the oil phantom. The coupled stack-up volume coil continues to demonstrate superior field homogeneity compared to the solenoid and equal gap coupled coils. In terms of E field efficiency, the coupled stack-up volume coil maintains significantly lower values, reinforcing its potential to reduce noise and enhance image quality in actual imaging scenarios.

**Figure 8.**
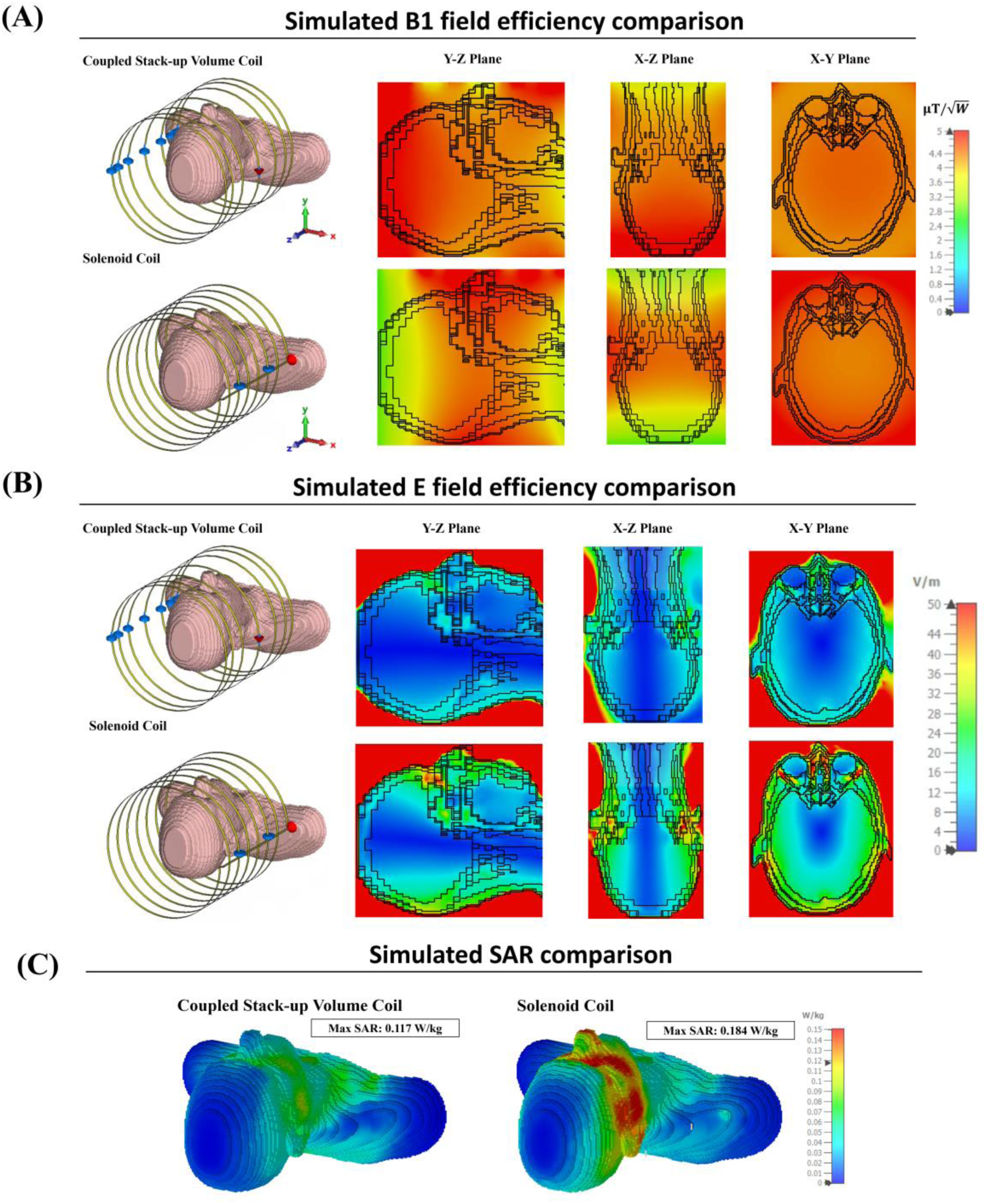
(A) Simulated loaded B1 field efficiency comparison with a CST Studio bio-model loaded into the coils. (B) Simulated loaded E field efficiency comparison with the bio-model. (C) Simulated SAR comparison between the three coils.

Moreover, the SAR comparison in Figure 8C highlights a critical advantage of the coupled stack-up volume coil: it exhibits significantly lower SAR levels compared to the solenoid coil. This suggests that the coupled stack-up volume coil not only offers better homogeneity and lower noise but also ensures safer operation by minimizing power deposition, making it particularly suitable for prolonged imaging sessions in clinical applications.

Figure 9 compares the simulated B1 field efficiency between the coupled stack-up coil and birdcage coil on three different planes with the B1 field efficiency distribution map. Table 1 also compares the relative standard deviation and average B1 field efficiency of the B1 field strength inside the phantom between the field generated by the coupled stack-up coil, solenoid coil, and birdcage coil. The result shows that the coupled stack-up coil has significantly higher B1 field efficiency and B1 field homogeneity compared with the birdcage coil. With an average of 10.82 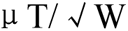 throughout the entire volume of the phantom, which serves as the region of interest (ROI), the B1 field efficiency of the coupled stack-up volume coil is 47.6% higher than the average B1 field efficiency of birdcage coil. As for homogeneity, the standard deviation of B1 field generated by the coupled stack-up volume coil is also 218.75% lower than that of the birdcage coil. Figure 10 shows the comparison of the simulated B1 field efficiency between the coupled stack-up volume coil and conventional birdcage coil with human head bio model as load. At around 21MHz, the B1 field distributions of both coils are not significantly affected by the load, with the B1 field distribution inside the human phantom remaining essentially consistent with that when an oil phantom is used inside the coil. The B1 field efficiency of the coupled stack-up volume coil still significantly exceeds that of the birdcage coil.

**Figure 9.**
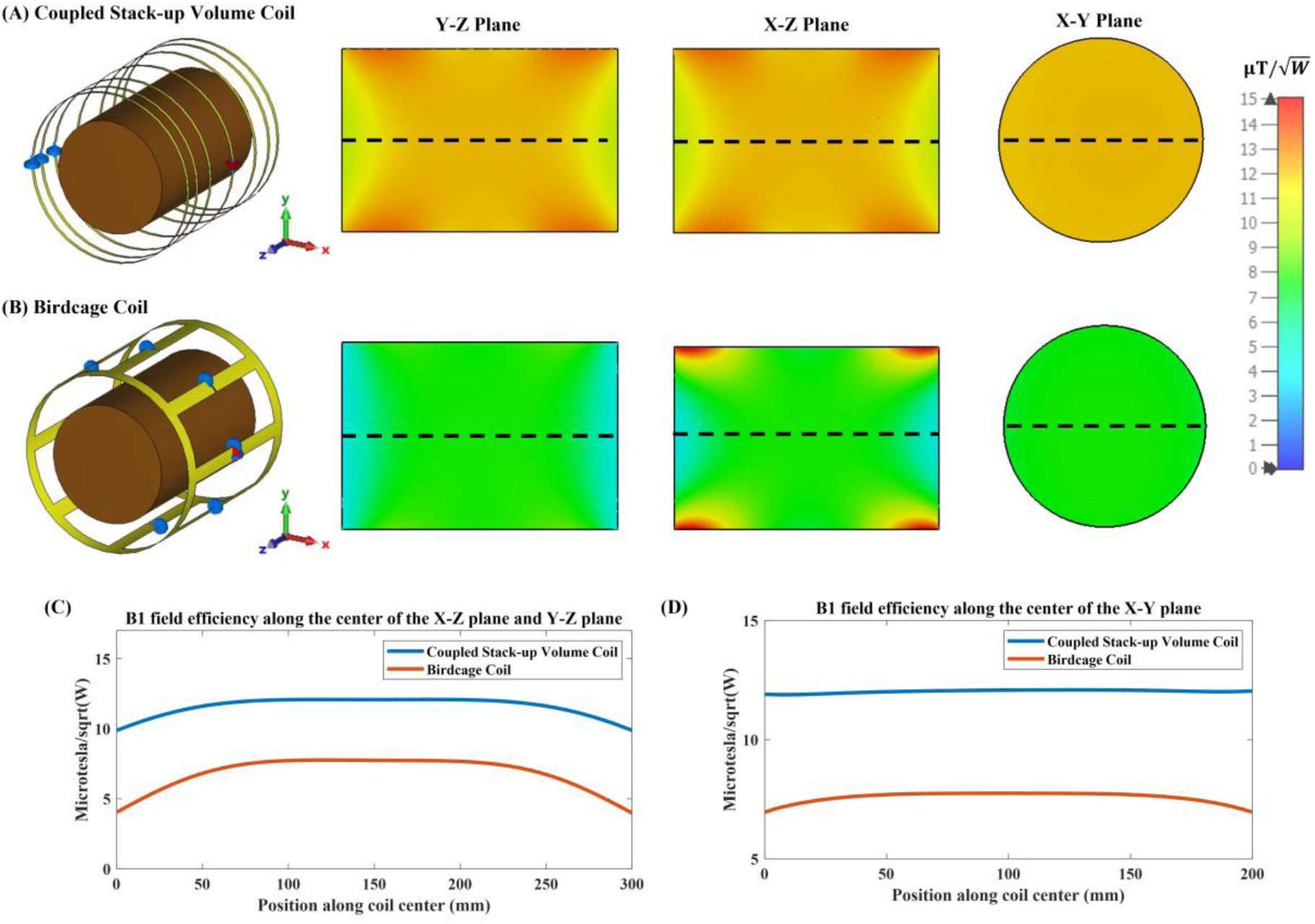
Simulated B1 efficiency and field distribution in three orthogonal planes: Comparison between the (A) proposed coupled stack-up volume coil and the (B) birdcage coil loaded with an oil phantom. (C)1-D profiles of the simulated B1 fields plotted along the axis of the coils, i.e. the dashed lines indicated in Y-Z plane and X-Z plane in inset (A) and (B). (D)1-D profiles of the simulated B1 fields of the coils plotted along the dashed lines shown in X-Y plane in inset (A) and (B).

**Figure 10.**
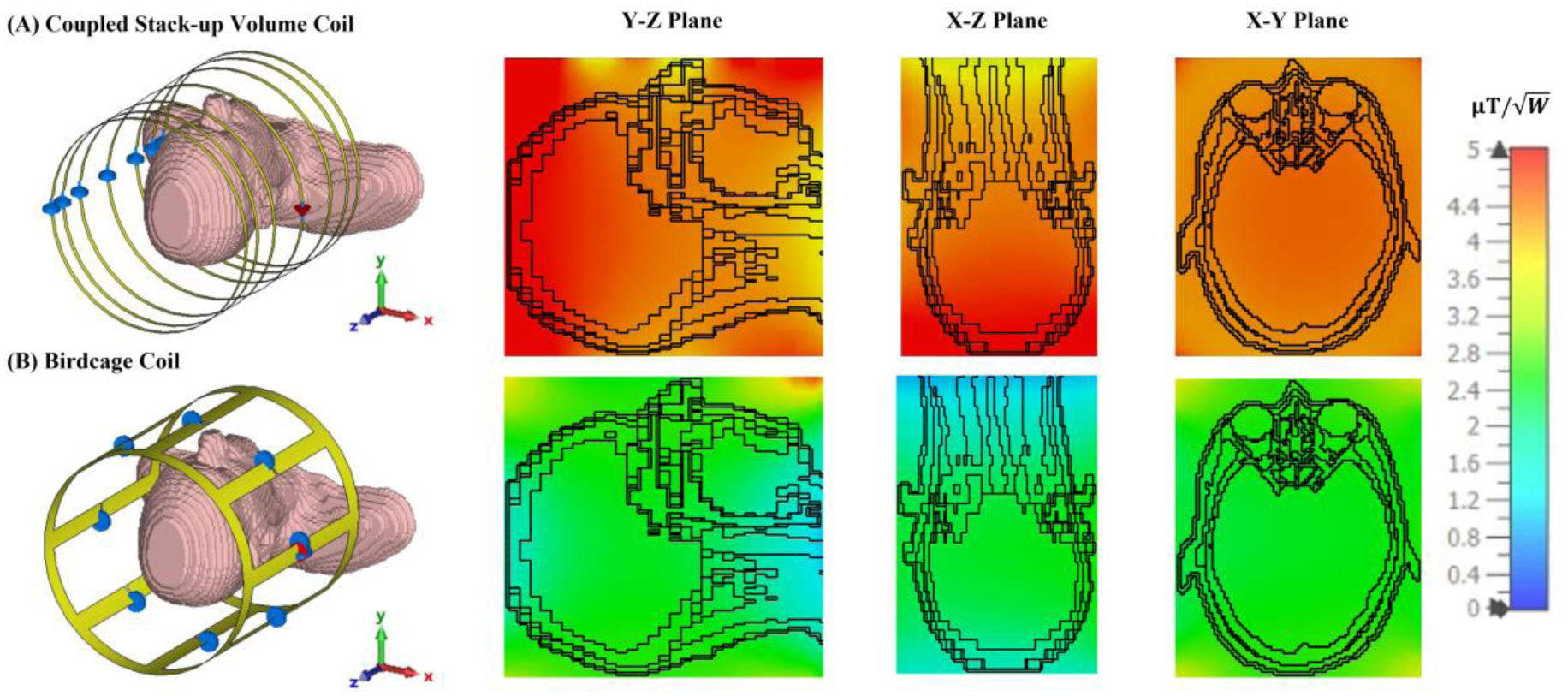
Simulated B1 efficiency and field distribution in three orthogonal planes: Comparison between the (A) proposed coupled stack-up volume coil and the (B) birdcage coil loaded with a human head phantom.

**Table 1.** Simulated average B1 field efficiency and standard deviation inside the phantom of coupled stack-up volume coil, solenoid coil, and birdcage coil. The average B field efficiency and relative standard deviation are calculated by the simulation result inside the phantom. Field efficiency is collected and analyzed in a 2.5 mm step size inside the phantom.

|  | Coupled Stack-up<br>Volume Coil | Solenoid Coil | Birdcage Coil |
| --- | --- | --- | --- |
| Average B Field<br>Efficiency ($\mu T/\sqrt{W}$) | 10.82 | 10.48 | 7.33 |
| Field Homogeneity<br>(Relative standard<br>deviation) | 4.81% | 10.02% | 15.30% |

Figure 11 compares the B1 field efficiency of the bench test model of the coupled stack-up volume coil and birdcage coil. The measured B1-field efficiency distribution is shown in Figures 9A and 9B. The measured magnetic field efficiency plot is consistent with the simulation results. Figures 9C and 9D show the B1 field efficiency plot at the center line along the X-Z plane, Y-Z plane, and X-Y plane. Not only does the coupled stack-up volume coil have higher B field efficiency, but the rate of decreasing of the B1 field from the center to the sides of the birdcage coil is much higher. The B field efficiency of the coupled stack-up volume coil, with the highest field efficiency of 11.48 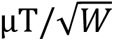, only reduces by 11.4% when reaching the edge of the coil with a minimum value of 10.20 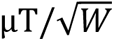. On the other side, the B1 field efficiency of the birdcage coil decreases by 49.20% from a maximum field efficiency of 7.30 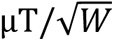 at the center to a minimum of 3.73 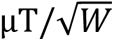 at two edges. The B1 field efficiency of the coupled stack-up volume coil is 57.30% higher compared with the birdcage coil. The measured result validates that the coupled stacked coil has a strong and homogeneous field within the imaging area compared with the birdcage coil.

**Figure 11.**
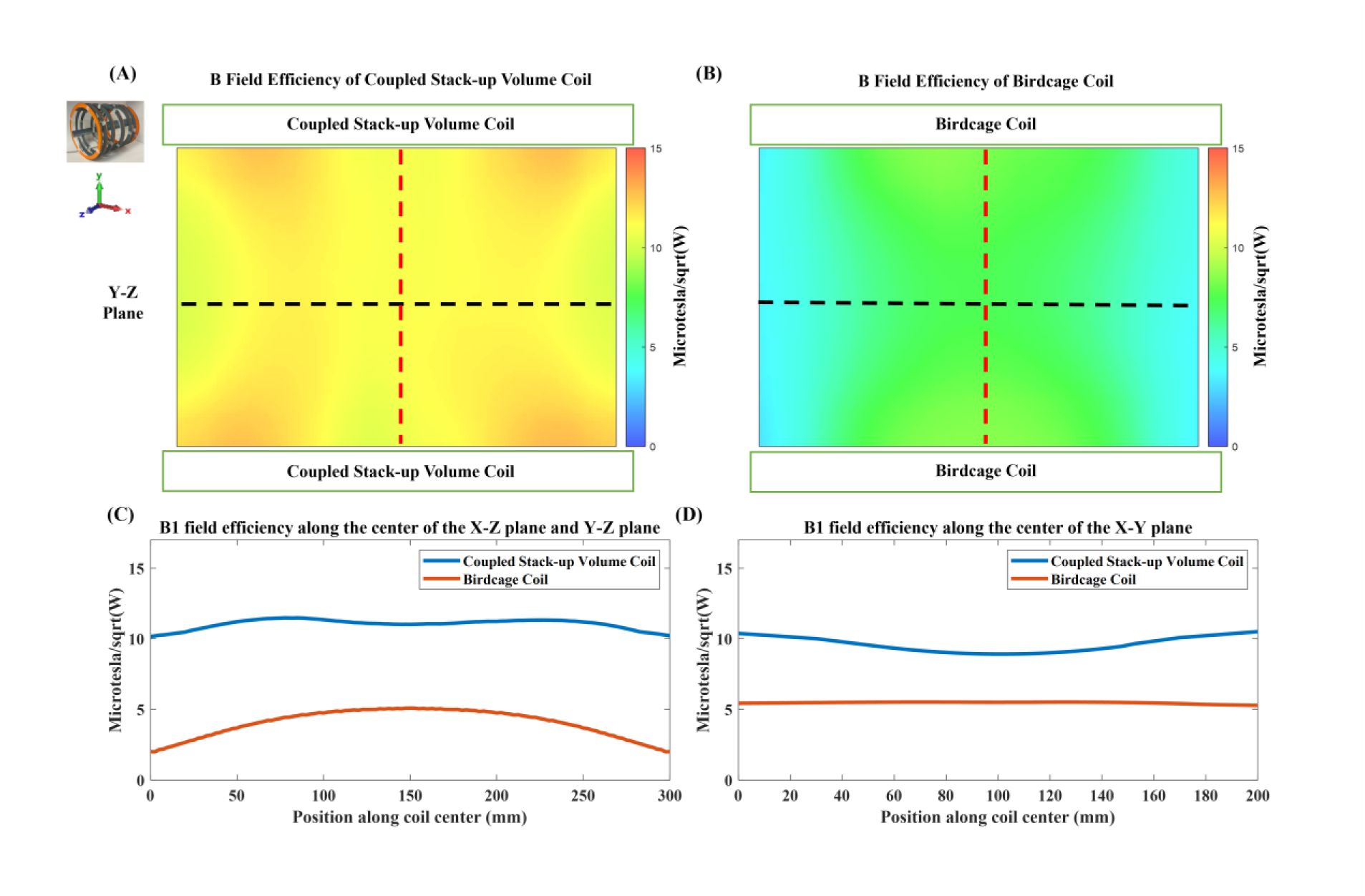
Measured unloaded B1 fields of the proposed coupled stack-up volume coil (A) and the same-sized birdcage coil (B). 1D profiles of B1 fields of the two coils plotted along the center line of the X-Z plane and Y-Z plane (black dashed lines in (A) and (B)) are shown in (C). 1D profiles of B1 fields of the two coils plotted along the center line of the X-Y plane (red dashed lines in (A) and (B)) are shown in (D). These results demonstrate the improved B1 efficiency and homogeneity of the coupled stack-up volume coil over the birdcage coil at 0.5T.

### 3.5 Effect of Increasing the Number of Rings

Figure 12 illustrates the impact of increasing the number of rings in the coupled stack-up volume coil on B1 field efficiency. Similar to the solenoid coil, the B1 field efficiency increases as the number of rings is increased. However, due to the width of the copper tape used to form the rings, it becomes increasingly challenging to find sufficient space to adjust the gaps between the rings. This limitation makes it more difficult to achieve a homogeneous field distribution, highlighting a trade-off between field efficiency and field homogeneity as the number of rings is increased.

**Figure 12.**
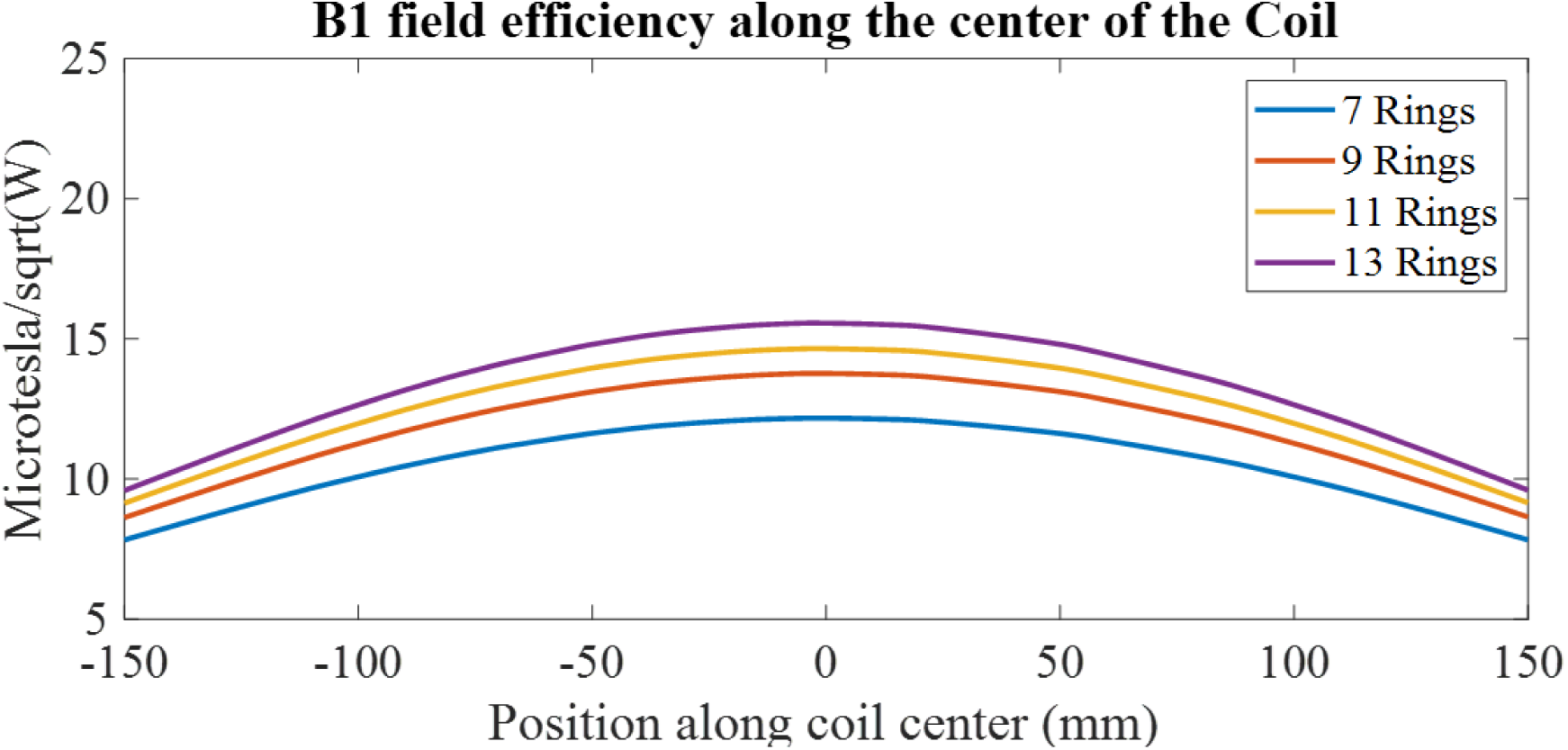
1-D profile comparison of the B1 field efficiency for the coupled stack-up volume coil with an increased number of rings.

## 4. Discussion

Critical to the success of the coupled stack-up volume coil design is the meticulous arrangement of its individual coils or resonant elements. The magnetic field strength at each coronal plane within the phantom should be most affected by the coil closest to it. By moving the coupled coil closer to the edge of the phantom where the B1 field strength is weaker, the local B1 field can be improved to match the B1 field strength at the center of the coil, thus improving the overall field homogeneity.

In the realm of low-field open MRI systems at 0.5 Tesla, the pursuit of enhanced image quality, diagnostic accuracy, and patient comfort has led to innovative approaches and technologies. This study has introduced the coupled stack-up volume coil, a novel RF coil design engineered to address the challenges inherent in 0.5T open MRI systems, particularly with respect to transmit/reception efficiency and field homogeneity. Through a research framework encompassing electromagnetic simulations and benchtop characterizations, we have illuminated the substantial advantages offered by this innovative coil design.

The adjustable nature of the coupled stack-up volume RF coil design could significantly enhance the versatility and performance of low-field MRI systems. By improving B1 field efficiency and homogeneity, this coil design addresses some of the inherent challenges associated with low-field imaging, such as lower signal-to-noise ratios. Moreover, the principles of this coil could be adapted to create flexible body coils, which are increasingly important in modern MRI applications. Flexible coils can conform better to the patient’s anatomy, leading to improved image quality and patient comfort. The ability to adjust the coil for different body parts would make it a versatile tool in clinical settings, particularly for imaging anatomically complex regions or for use in scenarios where patient movement is a concern. This adaptability could further extend the clinical applications of low-field MRI, making it a more viable option in various diagnostic scenarios.

Despite the advancements introduced by the coupled stack-up volume RF coil, several challenges remain. One of the primary challenges is the need to further optimize the coil for different body parts and imaging scenarios, particularly in the context of flexible designs. Additionally, while the current design demonstrates significant improvements in field efficiency and homogeneity, there is still room for further enhancement, particularly in reducing the complexity of the design without sacrificing performance. Future design approaches might explore the integration of advanced materials or novel coil geometries to further improve the coil’s adaptability and efficiency. Moreover, conducting extensive in vivo testing and developing more robust models for predicting coil performance across a range of conditions will be critical for advancing the clinical utility of these designs.

## 5. Conclusion

In conclusion, the coupled stack-up volume coil is successfully designed, constructed, and tested for low-field MR imaging. The proposed work represents a transformative development in the field of low-field MRI, particularly open MRI. Its innovative design, carefully arranged coil spacing, and optimized capacitance parameters converge to deliver a solution that outperforms the conventional birdcage coil in the aspects of B1 field efficiency, imaging coverage, and easy design and construction. It not only addresses the challenges posed by low-field MRI but also enhances its capabilities. The ability to achieve superior transmit/receive efficiency and field homogeneity positions this design as a promising avenue for advancing low-field MRI’s diagnostic precision and clinical utility.

## Data Availability

All data produced in the present study are available upon reasonable request to the authors

## Acknowledgments

This work is supported in part by the NIH under a BRP grant U01 EB023829 and by the State University of New York (SUNY) under SUNY Empire Innovation Professorship Award.

## Notes

### Competing Interest Statement

The authors have declared no competing interest.

